# Retrospective analysis of the clinical characteristics of *Candida auris* infection cases over a 10-year period

**DOI:** 10.1101/2020.05.19.20105817

**Authors:** Shan Hu, Yuehua Wang, Weiwei Jiang, Feilong Zhu, Yongqiang Quan, Guoming Zhang, Feng Gu, Bing Gu, Ying Yang

## Abstract

*Candida auris* is an emerging multidrug-resistant fungus with a high mortality rate. The first case of *Candida auris* infection was reported in 2009 and since then infections have been reported in nearly 40 countries. The fungus now represents a major global public health threat. We analyzed cases from the emergence of *Candida auris* infections up until the end of 2019.

PubMed and Web of Science databases were searched for all papers related to *Candida auris* infections up until 31 December 2019. We organized these data into the following categories: date of publication, patient age and gender, underlying diseases, risk factors for infection, patient mortality information, drug sensitivity information of *Candida auris* isolates, and genetic classification. The *χ*^2^ test was used to screen for factors that may affect patient mortality before logistic regression analysis was used to further assess the suspected influencing factors to determine if they represent independent factors for patient mortality.

Information pertaining to 542 patients was included. There were more male patients than female patients and the mortality rate was higher in males than females. A high proportion of patients were premature babies and elderly people. The proportions of patients with underlying diseases such as diabetes, kidney disease, and ear disease were also high. 65% of patients had a history in ICU and 60% were given broad-spectrum antibiotics. Logistic regression analysis revealed that kidney disease (P<0.05) and tumors (P<0.05) are independent factors that affect mortality in *Candida* auris-infected patients. Patients infected with echinocandin-resistant *Candida auris* ultimately die.

## Introduction

Since 2009 when the first *Candida auris* infection case was reported in Japan (1) until 31 December 2019, *Candida auris* infections have been reported in nearly 40 countries around the world (2). In April 2019, the New York Times reported a multidrug-resistant fungal infection outbreak in many parts of the United States. The causative agent of this outbreak was *Candida auris*; however, the aforementioned article referred to the infections as “mysterious infections” and deemed the outbreak an “urgent threat”. Nearly 50% of infected individuals died within 90 days and the outbreak subsequently attracted widespread global attention (3). Because of the associated high mortality rates, *Candida auris* can now be considered a major global public health threat (4).

Before 2009, *Candida auris* was thought to be a rarely observed microorganism and as a result it is now deemed an emerging human pathogen (5). *Candida auris* can colonize different sites, such as the skin, axilla, nose, and groin and is transmitted by contact or through feces. In addition, the pathogen can survive on inanimate object surfaces for more than 7 days (6, 7). Therefore, hospital beds, sphygmometers, thermometers, and other reusable equipment are potential infection sources for inpatients. This has resulted in relatively high rates of *Candida auris* transmission in inpatients, particularly in ICU rooms (8). This latter phenomenon has led to a vicious cycle of acquisition, transmission, and infection (9, 10). It is hard to eradicate *Candida auris* once it has colonized a patient and colonization of patients can last for 3 or more continuous months (11). Even though many countries have implemented infection prevention and control (IPC) strategies, *Candida auris* transmission is still a problem that warrants attention.

The appearance of multidrug resistance (MDR) in *Candida auris* is another problem that has attracted global attention in recent years (12). The sensitivity of many *Candida auris* isolates towards fluconazole has decreased and associated strains have developed varying degrees of resistance to other antifungal drugs (13). Public health guidelines recommend that echinocandins (micafungin, caspofungin, anidulafungin) should be used as first-line treatments for *Candida auris* infections (14, 15). However, the continuous evolution of *Candida auris* has resulted in echinocandin resistance. Indeed, the minimum inhibitory concentrations (MIC) of some isolates towards azoles, amphotericin B, and echinocandin have even increased (13, 16). Therefore, close attention is required in relation to the evolution of *Candida auris*.

Traditional identification techniques cannot be used to identify *Candida auris*. Therefore, the prevalence of *Candida auris* infections in the global population remains unknown. Furthermore, *Candida auris* has resulted in an “invisible pandemic” due to its broad nosocomial infection range (11). Thus, it is extremely important to comprehensively understand the characteristics of *Candida auris* itself along with the patients that the fungus infects. With this in mind, we collected papers from the last 10 years since the first report of *Candida auris* and extracted information pertaining to the patients’ age and gender, underlying disease, possible risk factors that may induce infection, genetic phenotype classification of *Candida auris* isolates, drug susceptibility information, and drug resistance loci. We compiled this information and conducted a statistical analysis to identify new infection-related characteristics. The mortality rate of *Candida auris* infection patients is an important factor that threatens global public health (4). Therefore, this study employed a conservative regression model to determine the effects of several factors on patient mortality. We believe that this analysis and summary of the relevant data will result in a greater public understanding of *Candida auris* infections, thereby enabling scientists to focus greater attention on the evolution of multidrug resistance in *Candida auris*.

## Methods

### Case inclusion

Authoritative literature search databases (PubMed and Web of Science) were used to conduct the literature search using the following search criteria: 1. Titles or abstracts containing the keyword *Candida auris;* 2. Time range with unlimited start date and an end date of 31 December 2019. Three hundred and sixty-five papers were identified through the literature search. Experimental study papers were excluded while case reports were selected for the analysis. A total of 76 papers on *Candida auris* case reports were included; these reports involved nearly 4000 patients (See Appendix 1).

The papers were organized and analyzed after case screening. Firstly, repeated cases were excluded: Suspected repetitive cases from the same year and country that were reported in some papers were excluded based on isolate number, detection time, and detection unit. Cases with incomplete information were excluded. As this study predominantly analyzed case characteristics, cases with incomplete information were excluded. The specific data of 542 detailed cases were included in the statistical analysis conducted as a part of this study.

### Data entry

The following information required for analysis was inputted:

1. Paper information (combined with data from the US Centers for Disease Control)^2^:
  1. Country of publication; (2) Year of publication;
2. Patient information;
  1. Basic information of patients: including gender, age, and whether patient is a premature infant.
  2. Underlying diseases of the patient: including common systemic diseases such as hypertension, diabetes, and hyperlipidemia; comorbid infections such as AIDS, syphilis, and cytomegalovirus (CMV) infection; solid organ diseases were classified based on the site, such as kidney disease, lung disease, and brain disease.
  3. Risk factors that may induce *Candida auris* infection: including use of central venous catheter, use of broad-spectrum antibiotics, parenteral nutrition, and intensive care unit admission.
  4. Patient mortality status: whether patient died and cause of death.
3. Isolate information:
  1. Drug sensitivity information: minimum inhibitory concentration (MIC) of common antifungal drugs and drug resistance loci;
  2. Genetic classification: Different isolates were classified based on genetic polymorphisms into: South Asia (clade I), East Asia (clade II), South Africa (clade III), and South America (clade IV) (17).

In order to ensure the accuracy of analysis, each inputted dataset required review by at least 3 members of the team before it was accepted. In this study, the Beijing Key Laboratory for Molecular Diagnostics of Infectious Diseases was deemed the leading site and it established a research team consisting of one clinician, two laboratory physicians, one medical mycology researcher, and one pharmacology researcher.

### Data organization and statistical analysis

In this study, underlying diseases predominantly included 3 categories encompassing systemic diseases, comorbid infection, and organ diseases; these were further divided into 50 subcategories. Suspected patients were not included in the statistics.

In order to avoid the repetition of statistical analyses pertaining to drug sensitivity data, the most recent data for an isolate was used when the same method was employed to analyze more than one drug sensitivity experiment. Interestingly, the MIC values for the most recent results generated from drug sensitivity experiments in all papers were the highest. When the broth method and standard commercial reagents were both used for drug sensitivity experiments for the same isolate, the drug sensitivity results obtained using standard commercial reagents were chosen for analysis because operator error tended to occur when the broth method was used. When different standard commercial drug sensitivity test systems (VITEK®2 and Sensititre® YeastOne® were used in this study) were used for the same isolate, the test results from the VITEK®2 system were selected as the inter-laboratory consistency of VITEK®2 was higher than that for Sensititre® YeastOne® (18).

The mortality rate of *Candida auris* infection is an important factor that threatens global public health. Hence, in this study we felt that it was important to screen for factors that affected patient mortality. The *χ*^2^ test was used to screen for single factors that might affect mortality. A p-value < 0.05 indicated that the suspected factor was significant. Significant suspected factors were included in multivariate analysis. If the number of significant suspected factors obtained during univariate analysis was less, the criteria for inclusion in the multivariate analysis were broadened to P < 0.1 (19, 20). For the multivariate analysis, we constructed a binary logistic regression model (21) to screen for independent influencing factors that affect the outcome (patient mortality). The outcome marker (death) was the dependent variable and all potential factors that affect patient mortality were independent variables. Potential factors that affect patient mortality included: gender, age, underlying disease, and drug resistance of the isolate. For influencing factors obtained by univariate analysis, only those with a partial regression coefficient of P < 0.05 in regression model analysis could significantly affect the outcome marker (patient mortality) (21).

## Results

### Initial infection reports in different countries

Since 2009 when *Candida auris* was first isolated from an auditory canal of a patient in Japan, infections have also been reported in South Korea (22), India (23), South Africa (24), and Kuwait (25). After 2016, the number of countries with first reports of *Candida auris* infections started to increase. In 2018, 14 countries reported *Candida auris* infections for the first time, this was the highest number of countries reporting infection for the first time in 10 years (shown in Figure 1).

**Figure 1:**
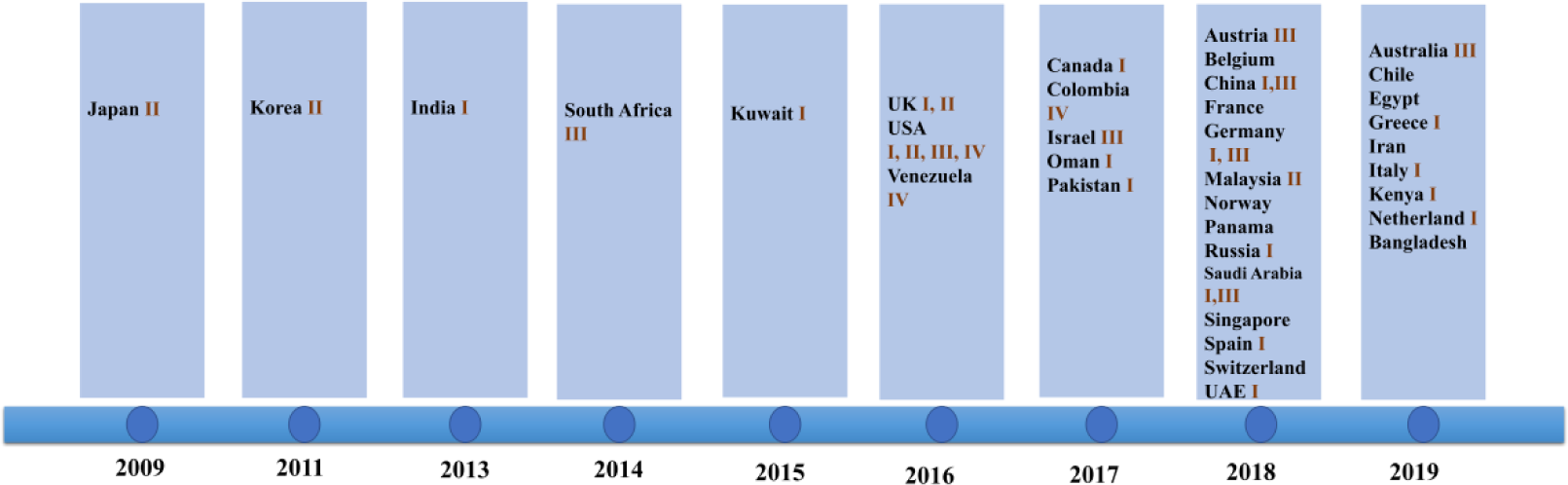
Date of first report and genetic classification for various countries (South Asia (clade I), East Asia (clade II), South Africa (clade III), and South America (clade IV))

### Genetic classification of isolates detected in various countries

The isolates detected by various countries were classified by whole genome sequencing data into South Asia strains (clade I), East Asia strains (clade II), South Africa strains (clade III), and South America strains (clade IV). The South Asia strains (clade I) were the most prevalent (17 countries), followed by the South Africa strains (clade III) which were observed in 8 countries. Only 4 and 3 countries reported the presence of the East Asia strains (clade II) and the South America strains (IV), respectively. Both South Asia and South Africa strains simultaneously occurred in China, Germany, UK, and Saudi Arabia. All 4 clades appeared in the US, a phenomenon that may be due to large population movements. In addition, an isolate identified in Iran in 2019 may represent a potential clade V strain (shown in Figure 1) (17).

### Patient gender and age distribution

For the included patients, gender was mentioned for 509 patients, of which 296 (58%) were male and 213 (42%) were females. The male to female ratio was around 3:2 and infection resulted in significant gender differences. This is shown in Figure 2a.

**Figure 2:**
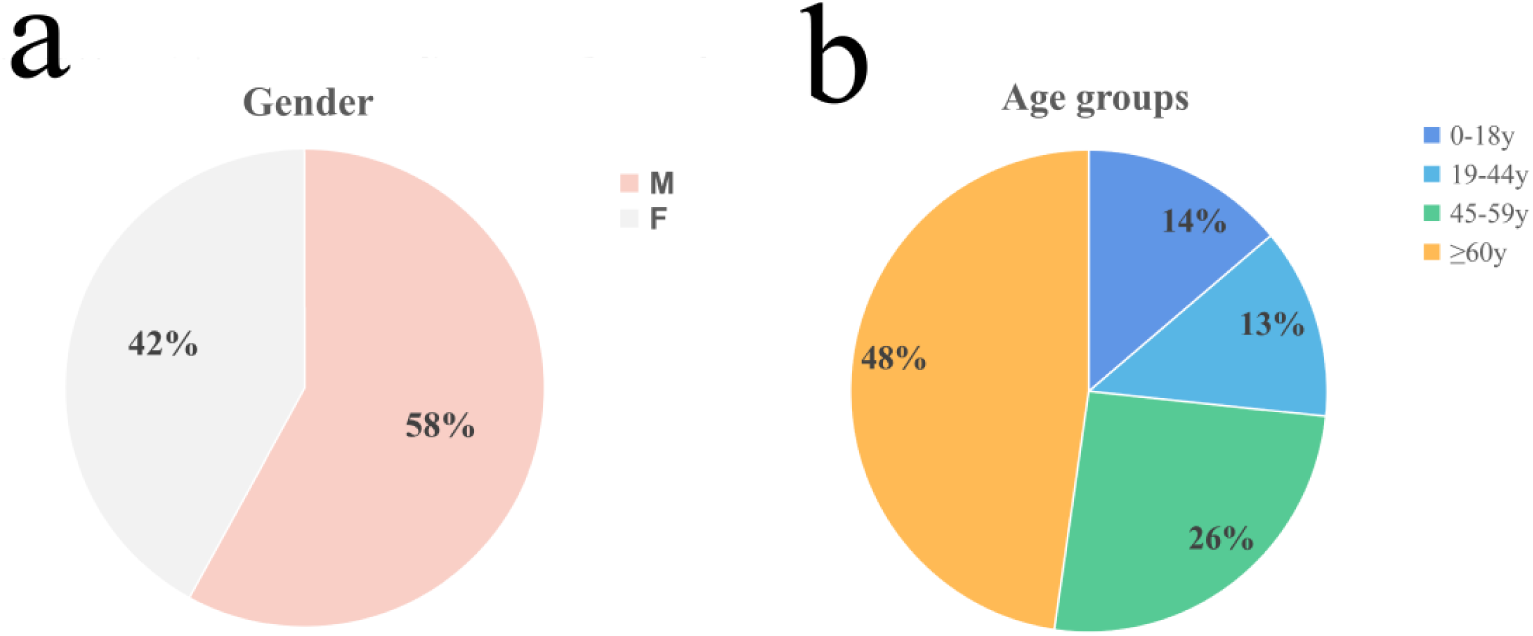
Gender and age distribution of patients. a). Gender distribution: males (58%) and females (42%). b). Age distribution: 0–18 years (14%), 19–44 years (13%), 45–60 years (26%), ≥60 years (48%).

The World Health Organization age group classification criteria (20) were used to divide patients into children (0–18 years), adolescents (19–44 years), middle-aged (45–59 years), and elderly (≥60 years). Figure 2b shows the age distribution of 195 cases in which age was mentioned. Most patients, approximately half of the population (n=93, 48%), were elderly. Fifteen of the patients were infants less than 1 month old; 10 of the 15 (2/3 of the infant population) patients were premature infants. Therefore, the possibility of *Candida auris* infection should be considered when premature infants present with infection symptoms due to unknown causes.

### Underlying disease and infected population

Underlying diseases were mentioned for 418 patients, of which the highest proportion of underlying diseases were diabetes, kidney disease (some have diabetic nephropathy), and ear disease. In addition, 12 patients were co-infected with HIV (as shown in Table 1).

**Table 1.**
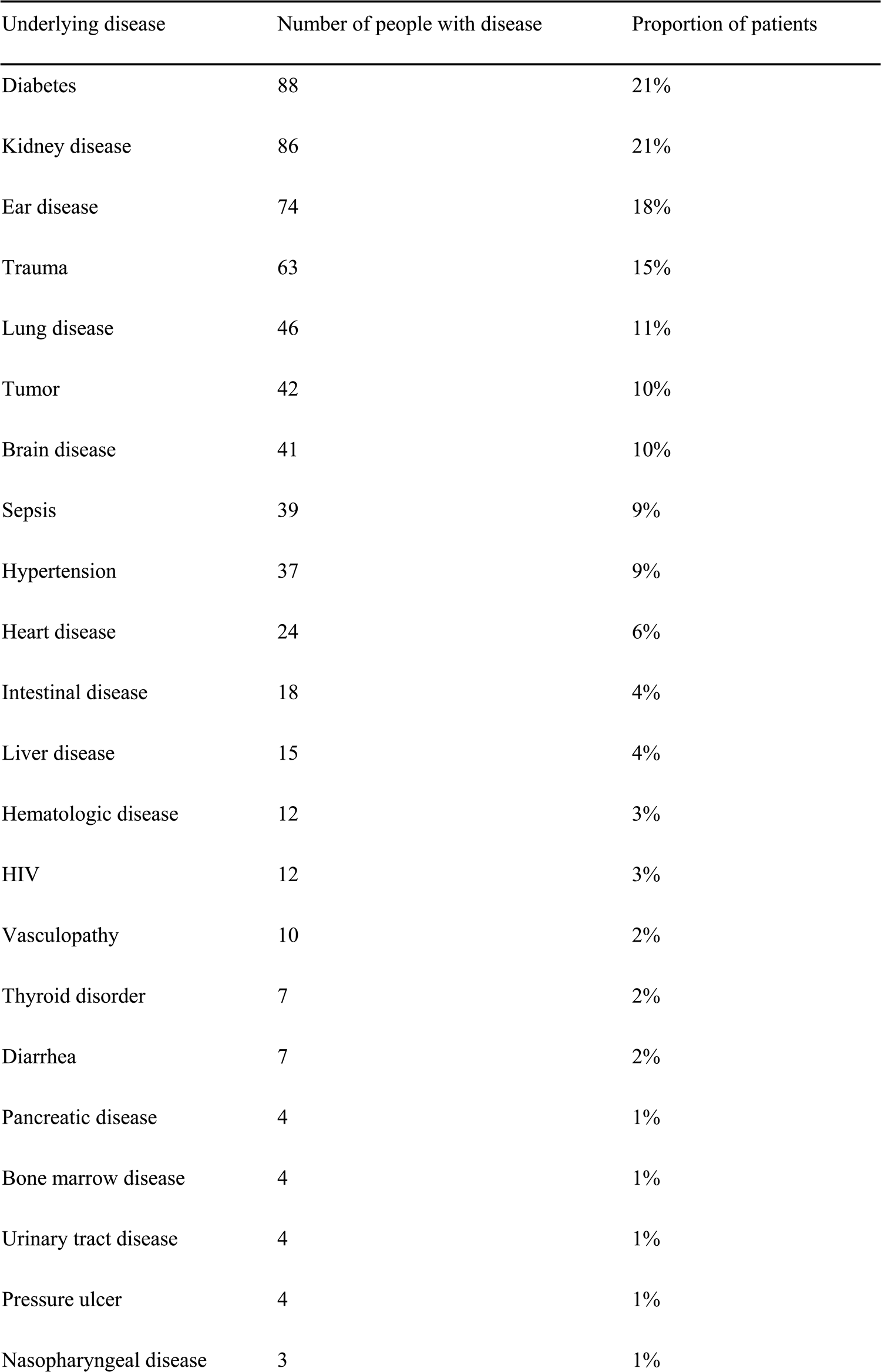
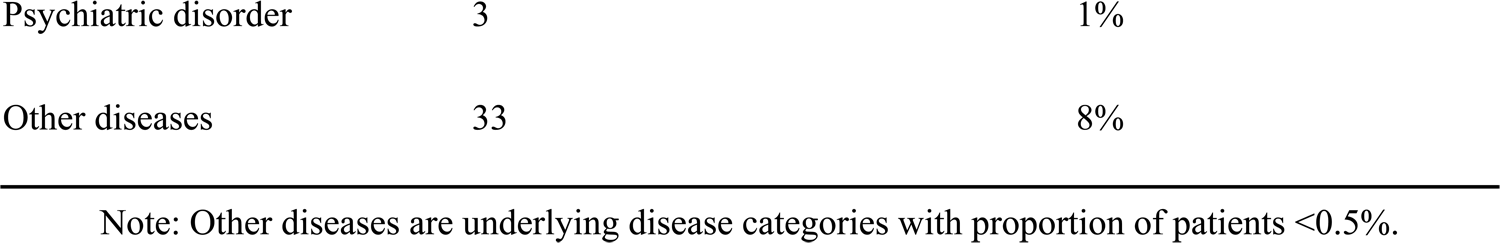
Underlying diseases in *Candida auris-infected* patients

### Risk factors that may lead to infection and infected population

Risk factors for infection were mentioned for 389 patients (Table 2). Greater than 60% of patients had an ICU treatment history and a history of broad-spectrum antibiotic use. These factors represent high risk factors for *Candida auris* infection and are consistent with the risk factors espoused by global experts in relation to *Candida auris* emergence and transmission (19,20).

**Table 2:**
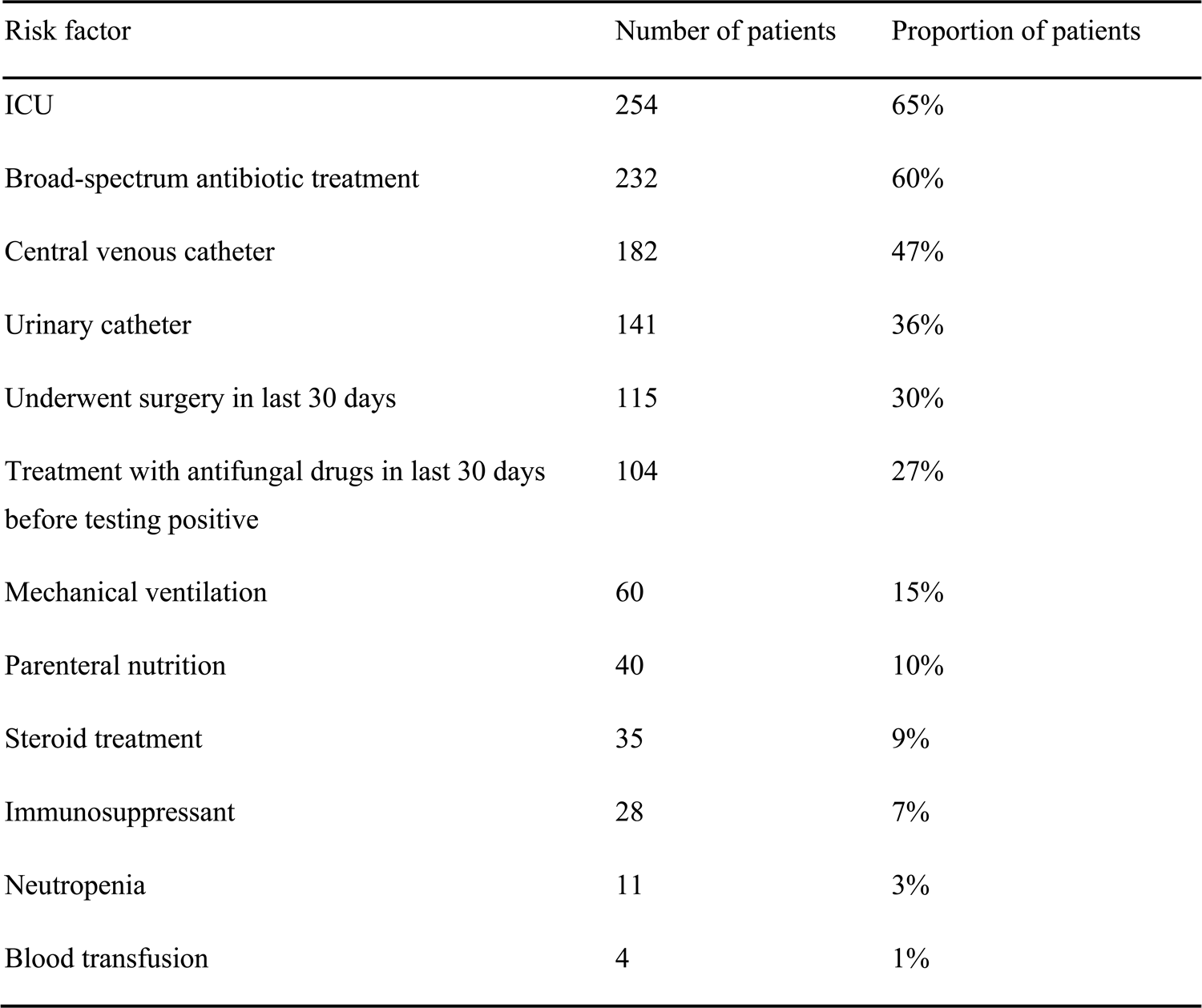
Risk factors involved in contact with *Candida auris* infected patients

### Drug-resistant *Candida auris* strains

Large differences in efficacy were observed for antifungal drugs in relation to the treatment of infection of different *Candida auris* isolates (13). The US Centers for Disease Control (CDC) defined conservative break points to determine whether *Candida auris* is resistant to antifungals: fluconazole (FLC): 32 μg /mL, amphotericin B (AMB): 2 μg/mL, micafungin (MFG): 4 μg/mL, and caspofungin (CAS): 2 μg/mL (2). In another study by the US CDC, break points for voriconazole (VRC): 2 μg/mL, anidulafungin (AFG): 4 μg/mL, and flucytosine (5-FC):128 μg/mL were established. A MIC < cutoff was defined as sensitive while a MIC ≥ cutoff was defined as resistant (25). Among the isolates that were included in the analysis, drug resistance information was available for 323 isolates; the proportion of isolates exhibiting FLC resistance was the highest (72%), while isolates exhibiting AMB (27%) and VRC (25%) resistance were the second and third most prevalent, respectively (Figure 3a). Among the VRC-resistant isolates, 18 Venezuelan isolates reported by Calvo et al. (7) and 8 USA isolates reported by Park et al (27). were completely resistant to VRC.

**Figure 3:**
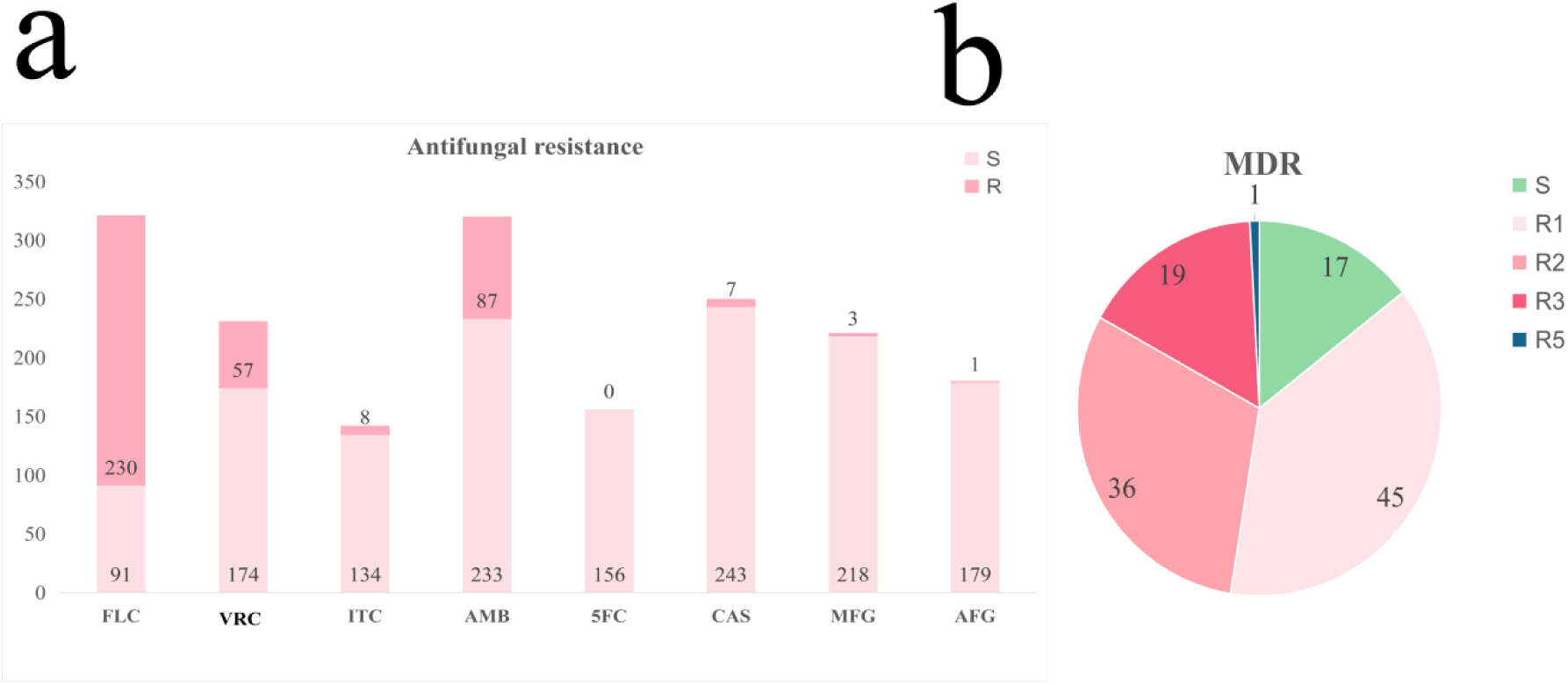
Drug resistance of *Candida auris*. a). Drug resistance status of *Candida auris* against 8 common antifungals. b). Drug resistance status of *Candida auris* isolates. MDR: multi-drug resistance. S: sensitive, R1-R5: drug resistance status

MIC information was accessible for 118 isolates in the literature and this information was used to determine drug resistance (Figure 3b). Among these isolates, 17 isolates (14%) were sensitive, 45 isolates (38%) were resistant to one antifungal, 36 isolates (31%) were resistant to two antifungals, 19 isolates (16%) were resistant to three antifungals, and 1 isolates (1%) was resistant to five antifungals. An analysis of these data revealed that there was no statistical correlation between the degree of multidrug resistance in *Candida auris* and patient mortality (Table 3). It should be noted that sensitive ones were isolated from 5 dead patients and the only sensitive isolate was from only a surviving patient. Only one isolate was resistant to 5 drugs (FLC, VRC, ITC, AMB, and CAS) and this isolate was obtained from a dead patient from Malaysia. Currently, echinocandin is the only first-line drug for *Candida auris* infection (14,15). When echinocandin resistance is present in the patient, the outcome is always death.

**Table 3:**
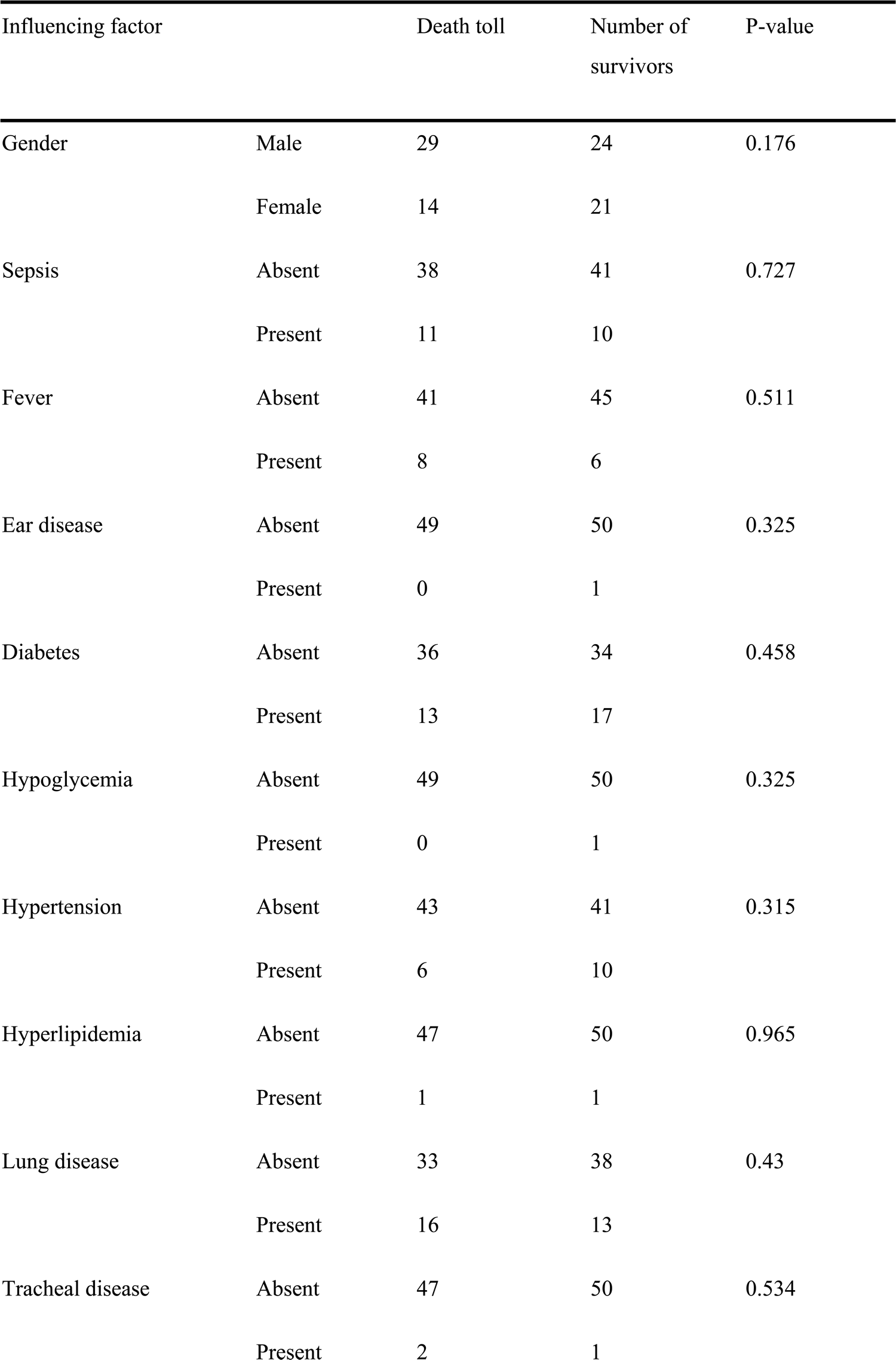
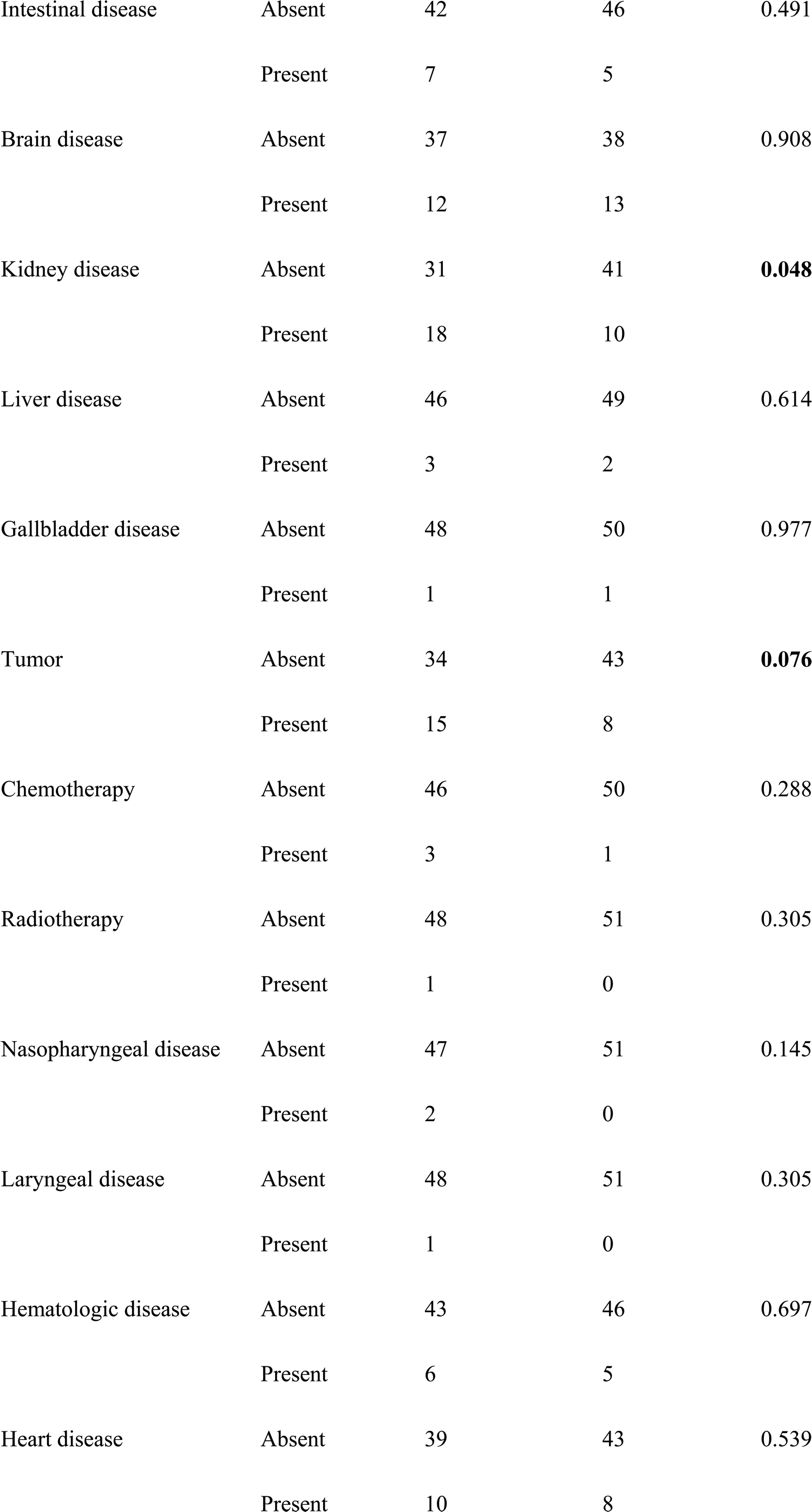
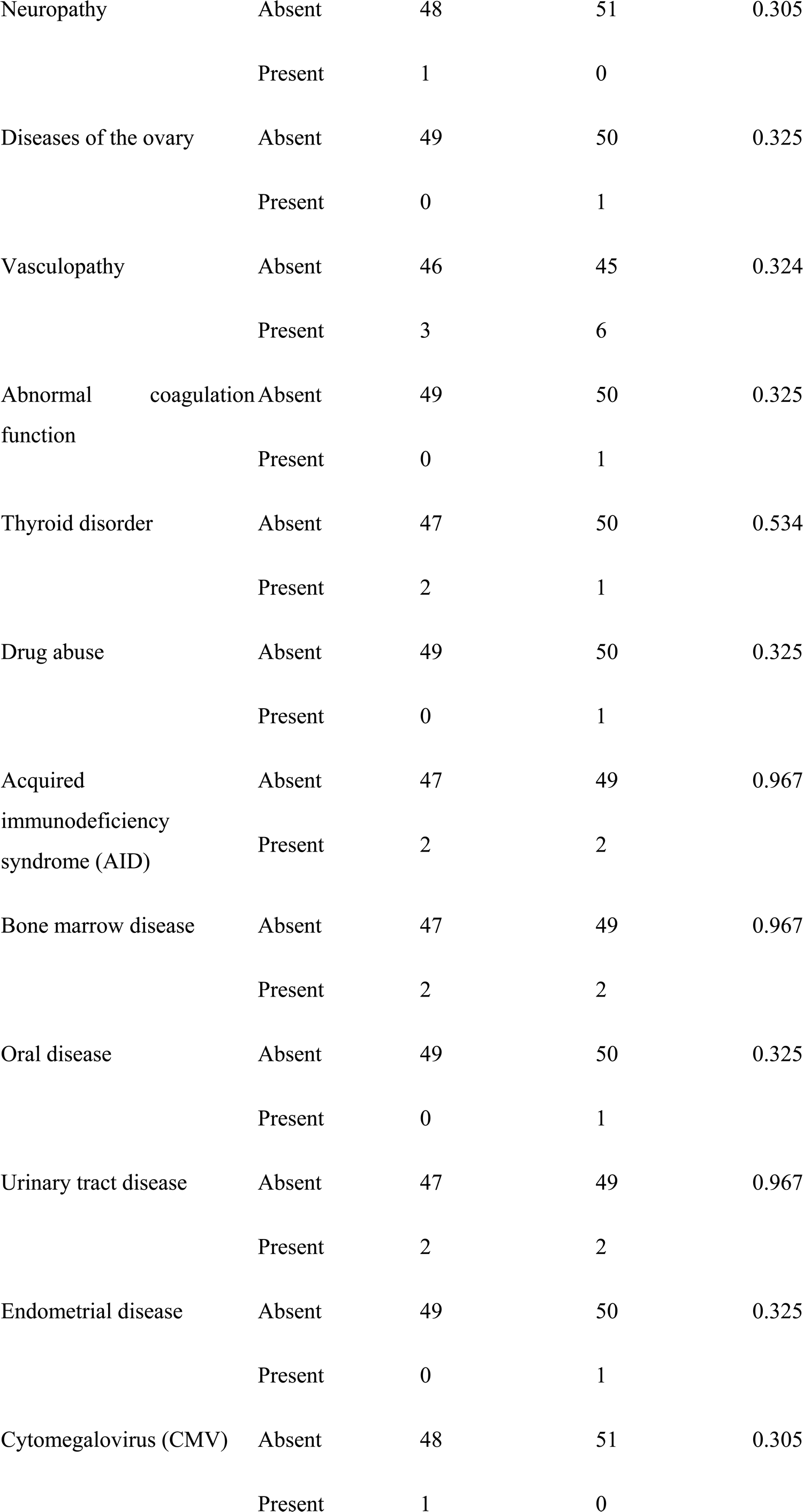
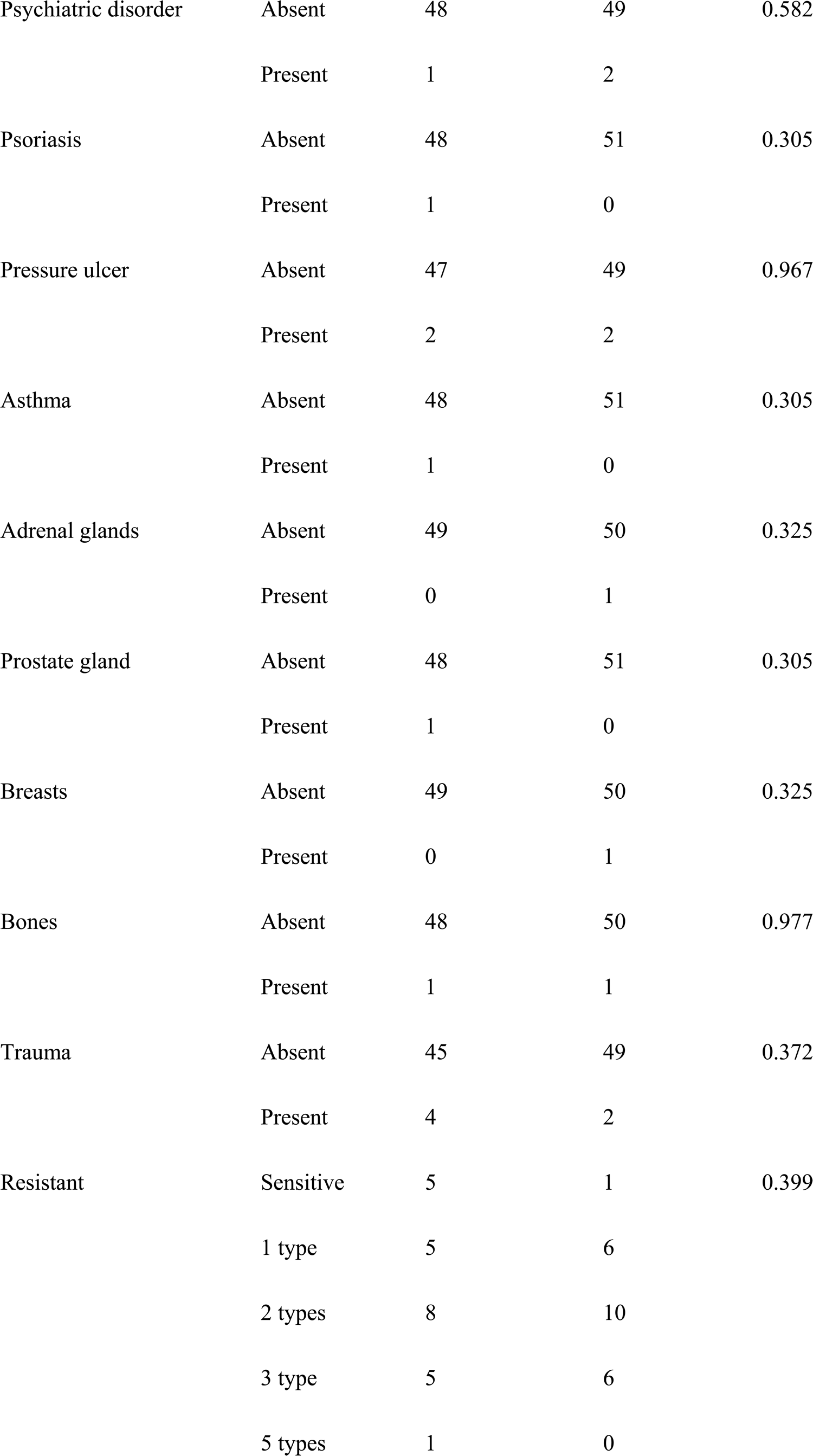
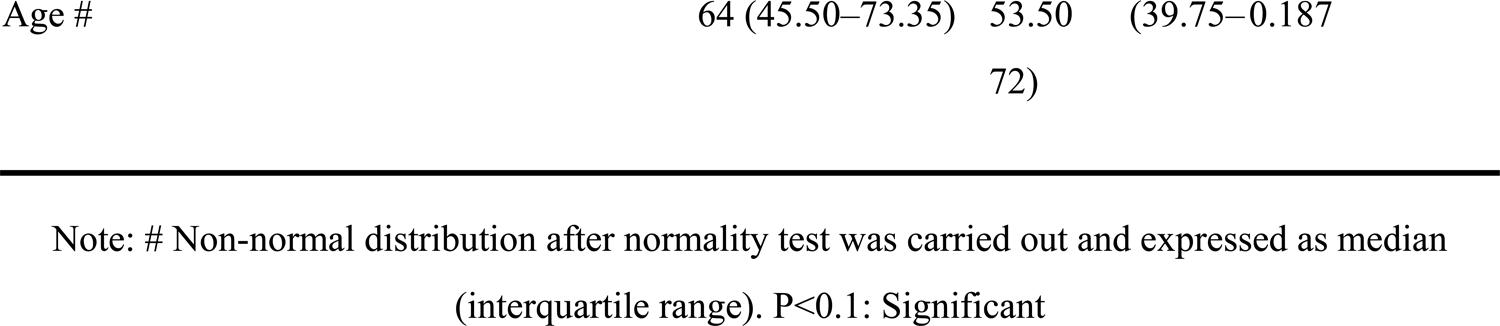
*χ*^2^ test screening of single factors that may affect patient mortality by category Influencing factor

### Drug resistance loci

Among the isolates reported to date, the major drug resistance loci for azoles are ERG11, CDR1, and MDR1 (27,28,29); the major drug resistance loci for AMB are ERG 2, 3, 5, 6, or 11 (30); and the major drug resistance locus for echinocandin is FKS11 (31,32). As part of a mechanistic analysis of drug resistance towards 5FC, Rhodes et al (16). sequenced the entire genome of 5FC-resistant *Candida auris* and found an amino acid substitution in F211I in the FUR1 gene. However, further studies are required to determine whether this FUR1 mutation is the cause of 5FC resistance in *Candida auris* (16).

### Mortality rate

A total of 218 cases reported whether patients died or survived upon discharge. Among the latter cases, 97 died and the mortality rate was 44%. Gender information was available for 88 dead patients, of which 29 out of 53 males died and the mortality rate was 53%; 14 out of 35 females died and the associated mortality rate was 40%; the mortality rate for males was slightly higher. Major causes of death that were mentioned included sepsis, septic shock, and multiorgan failure.

### Influencing factor for mortality

Among the patients included in the analyzed cases, the underlying disease of 100 patients corresponded to pre-discharge mortality and could be included in the analysis (in addition, many papers compiled patients’ information so that information could not be pertaining to each individual patient, which could not be included in the analysis). Gender, age, underlying disease, and drug resistance of isolates were used to screen for factors that affect mortality to discover influencing factors for mortality. The types of underlying diseases in dead and surviving patients were similar. First, the *χ*^2^ test was used to screen for single factors that may affect patient mortality by category (shown in Table 3). Among the factors screened, kidney disease had a P<0.05 and was analyzed by multivariate analysis. As the number of factors with P<0.05 was low, the inclusion criteria were relaxed to P<0.1 and tumor (P=0.076) was included in multivariate analysis as a suspected influencing factor (19,20).

Two suspected influencing factors (kidney disease and tumor) were selected based on the aforementioned univariate analysis and used for multivariate analysis. The outcome marker (mortality/survival) was used as a dependent variable and kidney disease and tumor were used as independent variables for logistic regression analysis (Table 4).

**Table 4:**
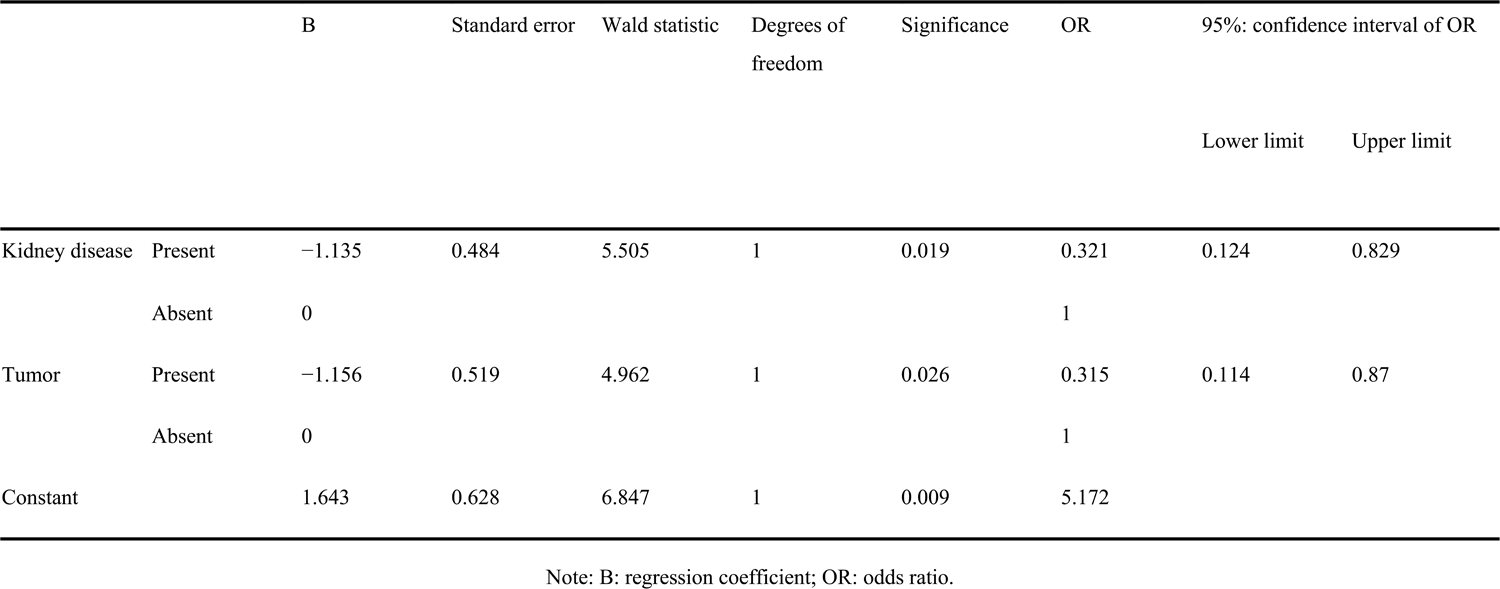
Multivariate analysis results

The following conclusions were ultimately drawn from the multivariate analysis:

1. Kidney disease is an independent influencing factor for the outcome marker (P=0.019<0.05) and the number of surviving patients with kidney disease was 32.1% of those without kidney disease. 2. Tumor is an independent influencing factor for the outcome marker (P=0.026<0.05) and the number of surviving patients with tumor is 31.5% of those without.

Based on the above conclusions, we selected kidney disease and tumor for prediction of patient outcomes and the prediction accuracy rate was 65.7%. The accuracy rate was deemed appropriate and the aforementioned conclusion had a good practical application value when the prediction accuracy rate was >60% (33).

## Discussion

*Candida auris* is an emerging pathogen and there is currently only a limited understanding of the mechanisms and influencing factors that underpin associated infections. Hence, further studies are required to help scientists better understand this pathogen. During the early stages of emergence of a pathogen, information release along with medical and public health awareness are extremely important.

In this analysis of collected literature, incomplete information was available for many patients. The main reason for these gaps in information relate to the fact that *Candida auris* isolates from these patients were not isolated during hospitalization but a few years later by public health institutions and research institutions during retrospective screening of samples. Furthermore, although *Candida auris* is difficult to culture many medical institutions still rely on culture to identify microorganisms (34). In addition, *Candida auris* is closely related to *Candida haemulonii* and *Candida pseudohaemulonii*. These three fungi are extremely close in phylogeny and it is difficult to distinguish them by phenotype. Thus, traditional identification methods such as Vitek 2 and API 20CAUX tend to wrongly identify *Candida auris* as these two closest relatives.^12^ Although the development of matrix-assisted laser desorption/ionization time-of-flight mass spectrometry (MALDI-TOF MS) (35) and sequencing (36) strategies have helped in the rapid and accurate diagnosis of *Candida auris*, most parts of the world do not have the infrastructure to carry out these techniques. In addition, it is likely that there is a large volume of unpublished data pertaining to *Candida auris* infections and the number of infected patients may be far higher than that reported in the literature. Therefore, this is an “invisible pandemic”.

In this retrospective analysis, we found that the male to female ratio for patients was nearly 3:2 and mortality data revealed that the male to female ratio for mortality was close to 4:3. Similar ratios have not been reported for other *Candida* infections (37). The proportion of *Candida auris* patients with diabetes, kidney disease, and ear disease is higher than that reported for other *Candida* infections. Diabetes is also a susceptibility factor for other candidiasis infections (20) but the high infection rates observed for kidney disease and ear disease are specific for *Candida auris*. Currently, almost all reported cases were from hospital wards. Jung et al. (21) reported that 78 out of 79 *Candida auris* infection patients had a history of consultation with the otolaryngology department; among these patients, 69 had ear disease and their symptoms were relatively mild. Therefore, otolaryngology outpatient departments should be highly vigilant for *Candida auris* infections. Among the *Candida auris* patient population, 21% had kidney disease; most of the latter patients had chronic kidney disease or nephrotic syndrome. More importantly, kidney disease is also an independent influencing factor that affects mortality in *Candida auris* infection patients. Therefore, nephrologists should be extremely vigilant for *Candida auris* infection. As the high infection rate in kidney disease patients is specific to *Candida auris* compared with other *Candida* species, nephrologists in many regions do not have high awareness for *Candida auris* infection. Many nephrologists do not completely understand *Candida auris*. Therefore, it is important that *Candida auris-related* information be disseminated to various medical units, particularly nephrology departments which can be extremely important in the early diagnosis of *Candida auris* infections. In addition, many *Candida auris* patients exhibit severe symptoms and although ICUs have been a focus of attention since *Candida auris* was first reported, overall vigilance in ICUs is still not very strong and there is a need to strengthen knowledge gaps pertaining to prevention, diagnosis, and treatment knowledge.

FLC exhibits broad-spectrum antifungal activity, good efficacy, a low incidence of adverse reactions, and appropriate plasma concentrations for long durations (38). Therefore, this drug is the most frequently utilized empirical therapy in patients suspected of fungal infection. *Candida auris* has a high FLC resistance rate and the overall drug resistance ratio has been as high as 72% over the last 10 years. Thus, FLC is not recommended as an empiric drug therapy when treatment is urgent and drug sensitivity results have yet to be released for patients who either have been infected with *Candida auris* or are suspected to have been infected with *Candida auris*. Clinicians need to develop an awareness when considering whether FLC should be used as empiric therapy in patients.

Although the data generated by this study reveal that there is no overall statistical correlation between the degree of multidrug resistance in *Candida auris* and patient mortality, we did identify some areas for attention after an in-depth analysis. Among the isolates for which patient mortality could be determined, sensitive ones were isolated from 5 dead patients and the only sensitive isolate was from only a surviving patient. We analyzed the possible reasons for the dearth of information pertaining to isolates causing mortality and speculated that virulence and mortality rates are higher when the *Candida auris* isolate is sensitive (39, 40). However, because sensitive isolates can be treated with antibiotics, clinical outcomes can be changed if the patients are diagnosed early and treated in a timely manner. Because candidiasis patients do not have apparent clinical characteristics at an early stage, early laboratory tests are key to reducing the mortality rate (11). Most Venezuelan and US patients exhibited VRC resistance. As these two countries are in the Americas, this phenomenon suggests that there is a relationship between antifungal treatment habits in different regions in the world.

In addition, among the acquired data, only one isolate that was resistant to 5 drugs was obtained and this was isolated from a Malaysian patient where the outcome was death. Therefore, when multidrug resistance in *Candida auris* gradually evolves, there will be situations where patients cannot be treated by drugs and death will result. However, because the outcome for patients infected with strains exhibiting only echinocandin resistance is death, vigilance is still required for instances where the fungi isolated from patients do not show severe drug resistance (e.g. resistance to 5 drugs). Echinocandin is currently the recommended treatment for *Candida auris*. However, if *Candida auris* gradually evolves, more and more strains will develop echinocandin resistance and this may lead to an increase in the mortality rate of patients with *Candida auris* infections. Although the β-glucan synthesis inhibitor SCY-078 (41) and other new drugs have shown some promise, they have not been widely used in clinical practice. Therefore, it is especially important to develop new drugs to combat high levels of echinocandin resistance. The appearance of antifungal drug resistance and the associated global range will continuously threaten global public health (42) and cannot be ignored.

There are some limitations pertaining to this study. As this is a retrospective analysis of published studies, there may be some original studies where the authors believed the associated conditions were not important and some of the influencing factors were not mentioned. Thus, these papers were not included in this current analysis. The stability of the drug sensitivity results from different locations may vary. However, this study is the largest retrospective analysis of Candida auris literature containing case data thus far. Additionally, as there was only a limited number of patients with certain diseases, there might be a selection bias in this study.

## Conclusion

This retrospective analysis provides us with a greater understanding of *Candida auris* along with some of the main characteristics associated with *Candida* auris-infected patients. For instance, the majority of the patients were male and had kidney disease or ear disease. It was also observed that kidney disease can affect patient mortality in infected individuals. In addition, an ever increasing number of multidrug resistant isolates have been identified and we should be vigilant in relation to the evolution of drug resistance in *Candida auris*. Moreover, it is important that we comprehensively improve diagnosis and treatment awareness for *Candida auris*. Furthermore, we should actively develop early diagnostic techniques along with safe and effective drugs for the treatment of *Candida auris* infections.

## Data Availability

Authoritative literature search databases (PubMed and Web of Science) were used to conduct the literature

https://www.ncbi.nlm.nih.gov/

## Acknowledgements

This research was supported by Ministry of science and technology of the People’s Republic of China (2018ZX10101003 and 2018ZX10712-001), Jiangsu Privincial Medical Talent (ZDRCA2016053), Six talent peaks project of Jiangsu Province (WSN-135)

## Research involving human participants and/or animals

Not applicable.

## Informed consent

Not applicable.

